# Genomic profiles of vaccine breakthrough SARS-CoV-2 strains from Odisha, India

**DOI:** 10.1101/2021.08.16.21261912

**Authors:** Arup Ghosh, Safal Walia, Roma Rattan, Amol Kanampalliwar, Atimukta Jha, Shifu Aggarwal, Sana Fatma, Niyati Das, Nirupama Chayani, Punit Prasad, Sunil K. Raghav, Ajay Parida

**Affiliations:** Institute of Life Sciences, Autonomous Institute under Department of Biotechnology, Government of India, Bhubaneswar, Odisha, India; SCB medical college and hospital, Cuttack, Odisha, India

**Author notes:** Equal contributors (Arup Ghosh, Safal Walia). **Correspondence:** Sunil Raghav (;), Ajay Parida (;).

**Keywords:** Vaccine Breakthrough, COVID-19, SARS-CoV-2, Variants of Concern

## Abstract

Vaccine breakthrough infections pose a vast challenge in the eradication of the COVID pandemic situation. Emerging SARS-CoV-2 variants of concern infecting the immunized individuals indicate an ongoing battle between host immunity and natural selection of the pathogen. Our report sheds light on the prominent SARS-CoV-2 variations observed in the isolates from AZD1222/Covishield and BBV152/Covaxin vaccinated subjects.

## Background

India has experienced 426,290 cumulative fatalities due to COVID-19 as of August 06, 2021 (https://covid19.who.int/table). Subsequent waves of SARS-CoV-2 infection with rapid resurgence in transmission and mortality deployed potential vaccine candidates to generate an effective immune response against SARS-CoV-2.

In India, five COVID-vaccines are authorized for emergency use, out of which the adenovirus-vector based vaccine from Oxford University and AstraZeneca UK marketed as Covishield, and the indigenous inactivated virus vaccine Covaxin by Bharat Biotech are majorly deployed through government and private healthcare centers. Both the vaccines pose tolerable safety outcomes and enhanced immune responses [1,2]. The emergence of novel variants of concern (VOC), novel spike (S) protein mutations leading to higher infectivity rate and vaccine escape poses threat to the vaccine induced protective responses. The Indian SARS-CoV-2 Genome Consortia (INSACOG) has identified presence of 4 major VOCs i.e., B.1.1.7, B.1.1.351, P.1, B.1.617.2 in India, that are associated with increased virulence and reduced efficacy of vaccines [3].

According to recent statistics from INSACOG portal (2021-08-06, http://clingen.igib.res.in/covid19genomes/) there is a drastic increase in frequency of B.1.617.2 lineage (Delta) from 8% to ∼75% after March, 2021 (Supplementary Image 1). Recent *in vitro* studies showed that sera from Pfizer or the AstraZeneca vaccinated individuals is less effective in neutralizing Delta variant in comparison to Alpha (B.1.1.7) [4]. In this study, we summarize 36 COVID vaccine breakthrough cases, which were SARS-CoV-2 RT-PCR positive despite evidence of an antibody response following vaccination.

## Methods

Data for this study were included from individuals who had been tested RT-PCR positive for SARS-CoV-2 after vaccination for either the BBV152 (COVAXIN) indigenous Indian vaccine or AZD1222 COVISHIELD (ChAdOx1) vaccine. Out of total 36 cases 34 received both the recommended doses of vaccine. All the cases were tested positive between the last week of March, 2021 and mid-June, 2021 after more than two weeks of vaccine second dose; while the remaining two patients tested positive within two weeks of their first dose of vaccination (Supplementary Figure 2).

Total RNA from collected nasopharyngeal swab samples was isolated using TANBead automated RNA extraction kit. The isolated RNA was subjected to qRT-PCR using Meril Covid-19 One Step RT-PCR Kit which includes amplification primers for viral ORF1ab and N gene for determining viral load by Ct values. Samples with Ct values < 35 were considered for the study.

Amplicon based dual indexed paired-end libraries for viral genome sequencing were prepared by COVIDSeq kit and sequenced using NextSeq-550 platform (Illumina). After demultiplexing, non-Host (Human) extracted reads using Kraken2 taxonomic classifier [5], were aligned using BWA against the Wuhan-Hu-1 (NC_045512.2) reference genome. Primer regions from the aligned sequences were trimmed using iVar followed by removal of duplicate reads. Single nucleotide variants and short INDELs were called using GATK4 Haplotypecaller using ploidy 1 followed by removal of low-quality variants (Quality by depth, QD < 5) by GATK VariantFiltration. Consensus sequences generated using BCFTOOL consensus and regions having no aligned reads were hard-masked. Clade and lineage of the viral genomes were determined using Nextclade (https://clades.nextstrain.org/) and PANGOLIN (pangoLEARN version 2021-07-28) respectively [6]. Rooted (root: NC_045512.2) phylogenetic tree of 549 sequences (522 other sequences collected in same timeframe from Odisha) was constructed using method described in [7]. All the calculations and statistical tests were performed using base R functions.

## Results

As a part of the regular COVID-19 genomic surveillance, we identified and sequenced 36 vaccine breakthrough infection cases. The study group consisted of 12 females, 24 males with age ranging from 23 to 65 years (median = 38.50, sd = 13.58) (Supplementary table 1). All subjects except two were fully immunized with two doses of either Covaxin/BBV1552(n=8) or AZD1222/Covishield (n=26). The interval between two vaccine doses ranged between 27 to 49 days with a median interval of 35 days (n = 34) and the onset of infection ranged between 6 to 98 days post vaccination (median = 74, sd = 25, n = 31). Out of 36 cases, 33 reported either single or a combination of common COVID-19 related symptoms i.e., fever, body pain, sore throat, one individual didn’t report any symptoms during sample collection and for the rest two samples the symptoms were unknown (Supplementary table 2). The specimens were collected between March 29 to June 15, 2021.

Out of the 36 cases, twenty nine were identified as variants of concern Delta (80%), two classified as Kappa (B.1.617.1) (∼6%), two as B.1 (∼6%), and one each in Delta (AY.3), B.1.575, B.1.629 lineages each. In COVAXIN break-through cases 9/9 classified as Delta variant, for COVISHIELD 20/27 were classified as Delta, 2/27 as Kappa, 2/27 as B.1, 1/27 as AY.3, 1/27 as B.1.575 and 1/27 as B.1.629 (Supplementary Figure 2, Supplementary table 2). When we looked for high frequency variants (present in > 20% of samples) in these two vaccination groups, COVAXIN cases contained an overall higher number of variants (n=42) in comparison to the COVISHILD group (n=28) (Figure 1 A, 1B). Although two of the cases were classified as B.1, we found both of them were classified as 21A (Delta) by Nextclade having S:D614G and S:P681R mutations along with other lineages (Supplementary table 3). We also found strong phylogenetic closeness of these B.1 lineages with Delta (B.1.617.2) variant cases collected in the same time frame (Figure 1C). Looking into Spike domain specific mutations, we observed emergence of EFR156G substitution (delE156,delF157, and R158G) in N-terminus domain, which is becoming a common trait of recently reported Delta (B.1.617.2) variants [8]. Interestingly, most of the breakthrough cases shared presence of S:L452R, S:T478K in receptor binding domain (RBD) along with S:D614G and S:P681R near S1-S2 furin cleavage site. All these spike variants have been proposed to be associated with increased infectivity through different mechanisms (Figure 1D) [9].

**Figure 1:**
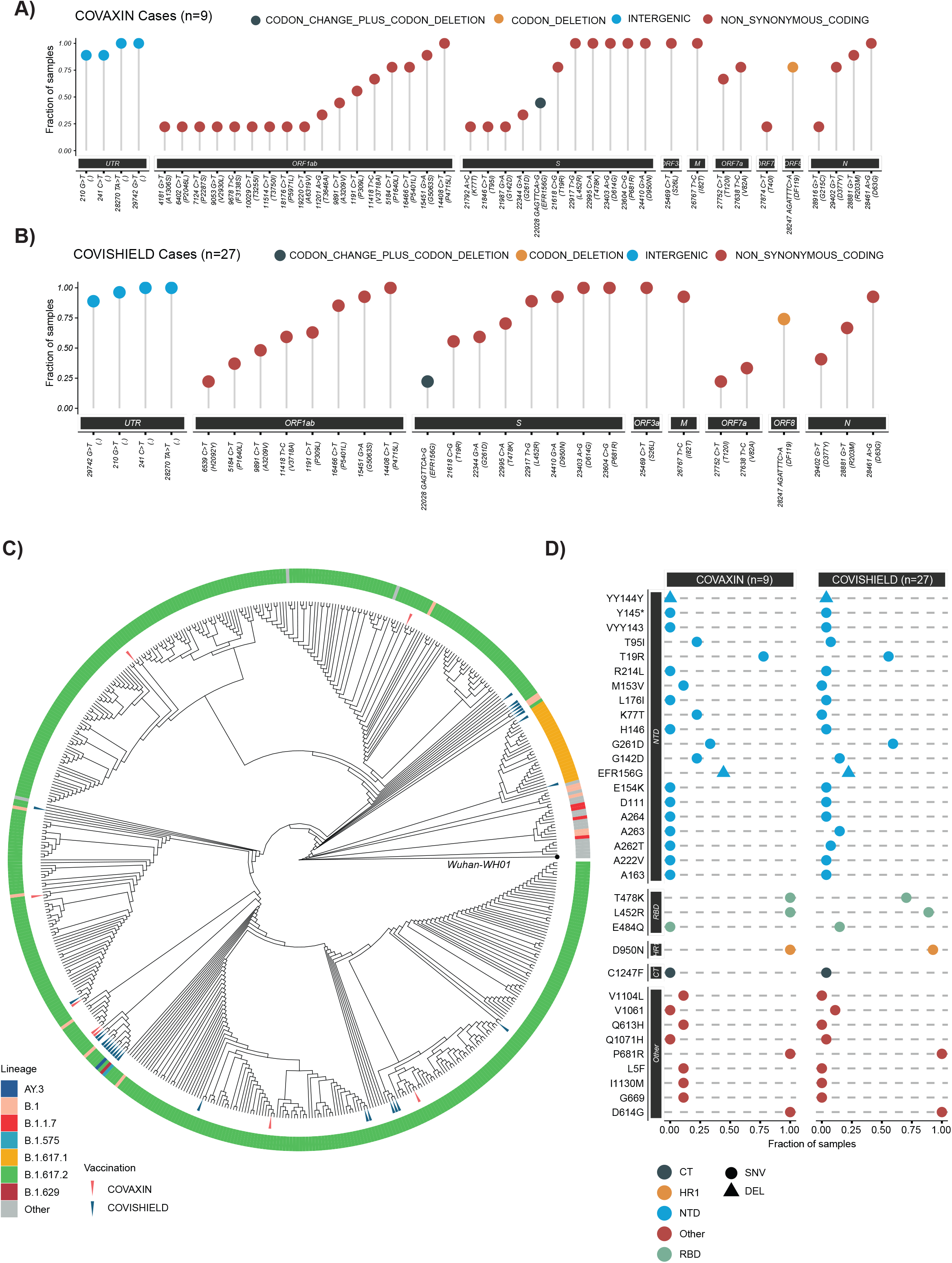
SARS-CoV-2 variants of concern and phylogenetic analysis of vaccine breakthrough cases; (A & B) Genome wide non-synonymous mutations present in high fraction (>20%) of samples, grouped by the vaccines, (C) Phylogenetic tree of our study genomes with other SARS-CoV-2 sequences (n=522) prevalent during the same time frame from Odisha, rooted using Wuhan Hu01 sequence as outgroup, and (D) Non-synonymous mutations observed in different domains of Spike protein grouped by vaccination type.

In the study group, 7 out of 36 patients reported comorbidities i.e., diabetes, blood pressure, rest 26 patients did not report any complications, disease history is unknown for three cases. Out of 36 patients only 1 patient was hospitalized with comorbidities, 23 recovered in home isolation, for 8 individuals the treatment status is unknown. Except for three unknown cases, the rest of the patients didn’t report any prior history of SARS-CoV-2 infection at the time of sample collection.

## Discussion

In this study we found that SARS-CoV-2 variant of concern Delta (B.1.617.2) is overrepresented in the vaccine break-through cases, which could be due to its higher prevalence as well during that period. Recent studies suggested that the effectiveness of BNT162b2 (Pfizer) and ChAdOx1 (AstraZeneca) has been reduced in comparison to Alpha (B.1.1.7) variant. Also the same has been observed in neutralizing capability of sera from fully vaccinated individuals [4,10]. One other *in vitro* study suggested that S:L452R and S:E484Q present in RDB domain of Spike protein are responsible for immune escape and thereby 3-6 fold decrease in neutralization capability of BNT162b2 sera [11].

Along with global reports of emerging variants of concerns and vaccine breakthrough infection cases now there are new threats emerging in India as well. Presence of two new Delta (AY.1, AY.2, AY.3) lineages has been observed in parallel with the Delta (B.1.617.2) that was first observed in India. Recent reports of breakthrough infection cases from India also have pointed out the predominance of Kappa (B.1.617.1) and Delta (B.1.617.2) variants [12].

Here we observed the co-occurrence of immune escape mutations and vaccine breakthrough infections despite the presence of COVID-specific immune response in hosts. Due to a small number of subjects and lack of more actionable clinically relevant information, we were not able to associate any direct causality for the breakthrough infections. But our findings are consistent with other recent reports showing the need for regular monitoring using genomics and immunological methods. Although we observed thirty-six breakthrough cases in a span of three months the number is very less in comparison to the total number of sequenced cases and the subject group mostly consisted of healthcare workers and thus the probability of observing breakthrough infection in a fully vaccinated population is very less. Continuing the virus genomic surveillance is necessary for devising new and improved intervention for COVID-19.

## Data Availability

ILS Intramural Grant.

## Acknowledgements

We would like to acknowledge the State Surveillance Officers (SSOs) of Odisha for their support in arranging the samples. Thanks to Dr. Andrew A. Lamare, Dr. Sudeep Jena and Mr. Rahul Biswal from SCB hospital, Cuttack for the samples and providing the metadata.

## Institutional Review Board approval

The study plan was approved by the Institute of Life Sciences Institutional Biosafety Committee (Ref no. V-122-MISC/2007-08/01) and the Institutional Human Ethical Committee (Ref no. 109/HEC/21). All necessary patient / participant consent has been obtained and the appropriate institutional forms have been archived.

