## Supplementary 1 for "Genomic profiles of vaccine breakthrough SARS-CoV-2 strains from Odisha, India"

**Supplementary Figure 1:** Variant of concern statistics (Data source: INSACOG Portal, Date:

2021-08-05,<https://clingen.igib.res.in/covid19genomes/>)

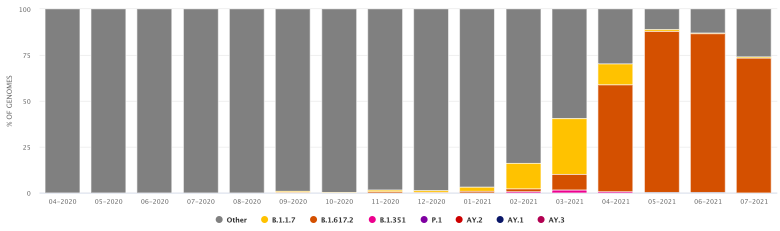

**Supplementary Table 1:** Summary of study groups

| **Age (years)** | **n=36** |
| --- | --- |
| Range | 23.00-65.00 |
| Median | 38.50 |
| Mean | 40.02 |
| SD | 13.58 |
| Q1 | 28.00 |
| Q2 | 38.50 |
| Q3 | 52.75 |
| Q4 | 65.00 |

| **Sex** | **n** |
| --- | --- |
| Female | 12 |
| Male | 24 |

|  | **Vaccine dose interval (days)** | **Time between vaccine dose and infection (days)** |
| --- | --- | --- |
| Range | 27.00-49.00 | 6.00-98.00 |
| Median | 35.00 | 76.00 |
| Mean | 34.24 | 67.97 |
| SD | 3.89 | 25.13 |
| Q1 | 35.00 | 66.50 |
| Q2 | 35.00 | 76.00 |
| Q3 | 35.00 | 81.50 |
| Q4 | 49.00 | 98.00 |

|  | **Ct (N gene)** | **Ct (ORF1ab)** |
| --- | --- | --- |
| Range | 14.00-30.77 | 15.00-30.16 |
| Median | 27.16 | 25.9 |
| Mean | 24.68 | 24.14 |
| SD | 5.23 | 4.73 |
| Q1 | 19.86 | 20.00 |
| Q2 | 27.16 | 25.90 |
| Q3 | 29.00 | 28.27 |
| Q4 | 30.77 | 30.16 |

* SD = Standard deviation, Q1= first quartile, Q2 = second quartile, Q3 = third quartile, Q4 = fourth quartile, Ct = Cycle threshold (RT-PCR)

**Supplementary Figure 2:** Cumulative number of breakthrough cases and their lineages

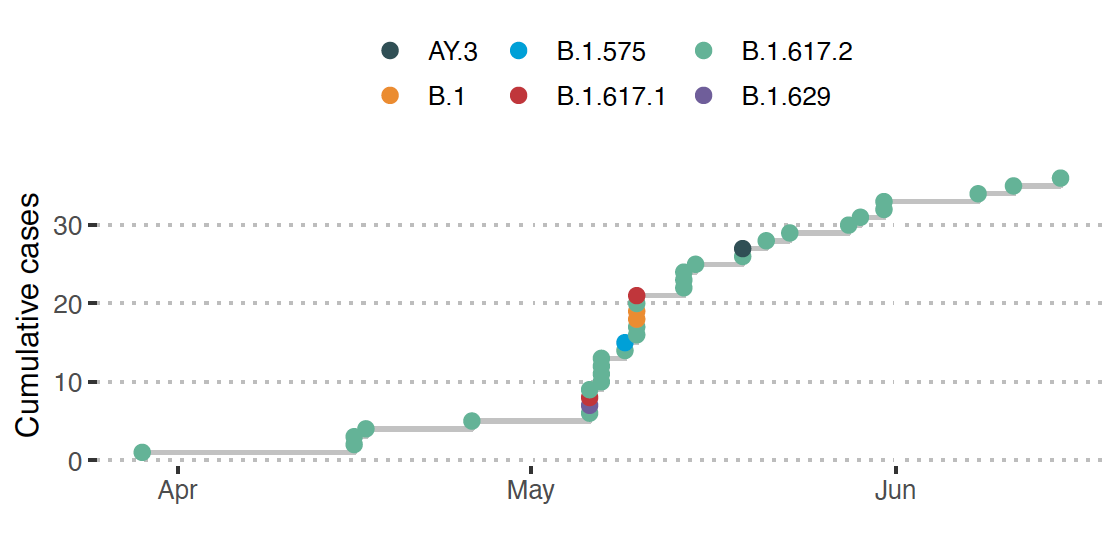

**Supplementary Table 2:** Case information

| **Patient** | **Age** | **Sex** | **Vaccine Type** | **Number of Dose** | **Last Dose to Collection Date (Days)** | **Ct (N gene)** | **Ct (ORF1ab gene)** | **Symptoms** | **Comorbidities** | **Clade (Nextclade)** | **Lineage(Pangolin)** |
| --- | --- | --- | --- | --- | --- | --- | --- | --- | --- | --- | --- |
| P001 | 31-35 | Female | COVAXIN | 2 | 18 | 18.00 | 19.00 | Fever,Body ache,Sore throat,Cough,Cold,Weakness | None | 21A (Delta) | B.1.617.2 |
| P002 | 26-30 | Male | COVAXIN | 1 | 15 | 14.00 | 15.00 | Body pain,Fever,Cold | None | 21A (Delta) | B.1.617.2 |
| P003 | 56-60 | Male | COVAXIN | 2 | 24 | 25.01 | 23.59 | Asymptomatic | Diabetes,Hypertension | 21A (Delta) | B.1.617.2 |
| P004 | 51-55 | Male | COVAXIN | 2 | 37 | 14.00 | 16.00 | Cold,Fever | Unknown | 21A (Delta) | B.1.617.2 |
| P005 | 26-30 | Male | COVAXIN | 2 | 76 | 26.00 | 26.00 | Sore throat | None | 21A (Delta) | B.1.617.2 |
| P006 | 26-30 | Female | COVAXIN | 2 | 21 | 17.00 | 17.00 | Sore throat | None | 21A (Delta) | B.1.617.2 |
| P007 | 56-60 | Male | COVISHIELD | 2 | 61 | 29.50 | 28.90 | Fever, Cold | Diabetes | 21A (Delta) | B.1.617.2 |
| P008 | 41-45 | Male | COVISHIELD | 2 | 62 | 28.60 | 25.80 | Fever, cough | Blood Pressure | 21A (Delta) | B.1.617.2 |
| P009 | 26-30 | Male | COVISHIELD | 2 | 71 | 30.10 | 29.60 | Fever | None | 21A (Delta) | B.1 |
| P010 | 61-65 | Male | COVISHIELD | 2 | 73 | 30.00 | 29.00 | fever, myalgia, headache, sore throat | Type 2 Diabetes Mellitus | 21A (Delta) | B.1.617.2 |
| P011 | 41-45 | Male | COVISHIELD | 2 | 73 | 26.00 | 29.00 | fever, myalgia, headache, sore throat | None | 21A (Delta) | B.1.629 |
| P012 | 46-50 | Female | COVISHIELD | 2 | 73 | 23.00 | 23.00 | fever, myalgia, headache, sore throat, cough >15days | None | 21B (Kappa) | B.1.617.1 |
| P013 | 36-40 | Male | COVISHIELD | 2 | 73 | 20.00 | 20.00 | fever, myalgia, headache, sore throat | None | 21A (Delta) | B.1.617.2 |
| P014 | 36-40 | Male | COVISHIELD | 2 | 74 | 29.00 | 27.00 | fever, myalgia, headache, sore throat | None | 21A (Delta) | B.1.617.2 |
| P015 | 26-30 | Male | COVISHIELD | 2 | 74 | 28.00 | 20.00 | fever, myalgia, headache, sore throat | None | 21A (Delta) | B.1.617.2 |
| P016 | 26-30 | Female | COVISHIELD | 2 | 76 | 28.00 | 29.00 | fever, myalgia, headache, sore throat | None | 21A (Delta) | B.1.617.2 |
| P017 | 26-30 | Male | COVISHIELD | 2 | 76 | 28.00 | 29.00 | fever, myalgia, headache, sore throat | None | 21A (Delta) | B.1.575 |
| P018 | 61-65 | Male | COVISHIELD | 2 | 77 | 28.00 | 27.00 | fever, myalgia, headache, sore throat, cough >15 days | None | 21A (Delta) | B.1 |
| P019 | 51-55 | Female | COVISHIELD | 2 | 77 | 29.00 | 28.00 | fever, myalgia, headache, sore throat | None | 21A (Delta) | B.1.617.2 |
| P020 | 61-65 | Male | COVISHIELD | 2 | 77 | 27.00 | 26.00 | fever, myalgia, headache, sore throat | Cancer | 20A | B.1.617.1 |
| P021 | 46-50 | Male | COVISHIELD | 1 | 6 | 17.54 | 19.05 | Unknown | Unknown | 21A (Delta) | B.1.617.2 |
| P022 | 56-60 | Male | COVISHIELD | 2 | 98 | 23.79 | 21.38 | fever, myalgia, headache, sore throat | None | 21A (Delta) | B.1.617.2 |
| P023 | 21-25 | Female | COVISHIELD | 2 | 98 | 30.77 | 30.16 | fever, myalgia, headache, sore throat | None | 21A (Delta) | B.1.617.2 |
| P024 | 21-25 | Male | COVISHIELD | 2 | 95 | 16.18 | 15.09 | fever, myalgia, headache, sore throat | None | 21A (Delta) | B.1.617.2 |
| P025 | 26-30 | Male | COVISHIELD | 2 | 88 | 28.00 | 27.00 | fever, myalgia, headache, sore throat | None | 21A (Delta) | B.1.617.2 |
| P026 | 26-30 | Female | COVISHIELD | 2 | 86 | 29.25 | 28.40 | fever, myalgia, headache, sore throat | None | 21A (Delta) | B.1.617.2 |
| P027 | 56-60 | Male | COVISHIELD | 2 | 86 | 29.44 | 29.43 | fever, myalgia, headache, sore throat | None | 21A (Delta) | AY.3 |
| P028 | 26-30 | Female | COVISHIELD | 2 | 82 | 29.29 | 28.22 | fever, myalgia, headache, sore throat | None | 21A (Delta) | B.1.617.2 |
| P029 | 21-25 | Male | COVISHIELD | 2 | 81 | 23.00 | 22.00 | fever, myalgia, headache, sore throat, cough >15 days | None | 21A (Delta) | B.1.617.2 |
| P030 | 26-30 | Female | COVAXIN | 2 | 74 | 16.67 | 17.27 | Fever,Body pain,Cold | Low Blood Pressure | 21A (Delta) | B.1.617.2 |
| P031 | 46-50 | Male | COVAXIN | 2 | 35 | 19.45 | 20.33 | Fever,Body pain,Cough | Diabetes, Hypertension | 21A (Delta) | B.1.617.2 |
| P032 | 51-55 | Female | COVAXIN | 2 | NA | 18.00 | 20.00 | Unknown | Unknown | 21A (Delta) | B.1.617.2 |
| P033 | 41-45 | Male | COVISHIELD | 2 | 96 | 27.33 | 27.37 | fever, myalgia, headache, sore throat, cough >15 days | None | 21A (Delta) | B.1.617.2 |
| P034 | 36-40 | Male | COVISHIELD | 2 | 89 | 27.72 | 24.37 | fever, myalgia, headache, sore throat | None | 21A (Delta) | B.1.617.2 |
| P035 | 36-40 | Female | COVISHIELD | 2 | 79 | 29.36 | 28.00 | fever, myalgia, headache, sore throat, cough >15 days | None | 21A (Delta) | B.1.617.2 |
| P036 | 26-30 | Female | COVISHIELD | 2 | 78 | 24.37 | 23.18 | fever, myalgia, headache, sore throat | None | 21A (Delta) | B.1.617.2 |

* Ct = Cycle threshold (RT-PCR)

**Supplementary Table 3:** Spike domain variants

| **Amino Acid Change** | **Effect** | **Region Name** | **COVAXIN count** | **COVISHIELD count** |
| --- | --- | --- | --- | --- |
| L5F | NON_SYNONYMOUS_CODING | Other | 1/9 | 0/27 |
| T19R | NON_SYNONYMOUS_CODING | NTD | 7/9 | 15/27 |
| K77T | NON_SYNONYMOUS_CODING | NTD | 2/9 | 0/27 |
| T95I | NON_SYNONYMOUS_CODING | NTD | 2/9 | 2/27 |
| D111 | SYNONYMOUS_CODING | NTD | 0/9 | 1/27 |
| G142D | NON_SYNONYMOUS_CODING | NTD | 2/9 | 4/27 |
| VYY143 | FRAME_SHIFT | NTD | 0/9 | 1/27 |
| YY144Y | CODON_CHANGE_PLUS_CODON_DELETION | NTD | 0/9 | 1/27 |
| Y145* | STOP_GAINED | NTD | 0/9 | 1/27 |
| H146 | SYNONYMOUS_CODING | NTD | 0/9 | 1/27 |
| M153V | NON_SYNONYMOUS_CODING | NTD | 1/9 | 0/27 |
| E154K | NON_SYNONYMOUS_CODING | NTD | 0/9 | 1/27 |
| EFR156G | CODON_CHANGE_PLUS_CODON_DELETION | NTD | 4/9 | 6/27 |
| A163 | SYNONYMOUS_CODING | NTD | 0/9 | 1/27 |
| L176I | NON_SYNONYMOUS_CODING | NTD | 0/9 | 1/27 |
| R214L | NON_SYNONYMOUS_CODING | NTD | 0/9 | 1/27 |
| A222V | NON_SYNONYMOUS_CODING | NTD | 0/9 | 1/27 |
| G261D | NON_SYNONYMOUS_CODING | NTD | 3/9 | 16/27 |
| A262T | NON_SYNONYMOUS_CODING | NTD | 0/9 | 2/27 |
| A263 | SYNONYMOUS_CODING | NTD | 0/9 | 4/27 |
| A264 | FRAME_SHIFT | NTD | 0/9 | 1/27 |
| L452R | NON_SYNONYMOUS_CODING | RBD | 9/9 | 24/27 |
| T478K | NON_SYNONYMOUS_CODING | RBD | 9/9 | 19/27 |
| E484Q | NON_SYNONYMOUS_CODING | RBD | 0/9 | 4/27 |
| Q613H | NON_SYNONYMOUS_CODING | Other | 1/9 | 0/27 |
| D614G | NON_SYNONYMOUS_CODING | Other | 9/9 | 27/27 |
| G669 | SYNONYMOUS_CODING | Other | 1/9 | 0/27 |
| P681R | NON_SYNONYMOUS_CODING | Other | 9/9 | 27/27 |
| D950N | NON_SYNONYMOUS_CODING | HR1 | 9/9 | 25/27 |
| V1061 | SYNONYMOUS_CODING | Other | 0/9 | 3/27 |
| Q1071H | NON_SYNONYMOUS_CODING | Other | 0/9 | 1/27 |
| V1104L | NON_SYNONYMOUS_CODING | Other | 1/9 | 0/27 |
| I1130M | NON_SYNONYMOUS_CODING | Other | 1/9 | 0/27 |
| C1247F | NON_SYNONYMOUS_CODING | CT | 0/9 | 1/27 |
